# Coronametrics: The UK turns the corner

**DOI:** 10.1101/2020.04.17.20069278

**Authors:** Adam Goliński, Peter Spencer

## Abstract

* There are many ways of analyzing the progress of an epidemic, but when it comes to short term forecasting, it is very hard to beat a simple time series regression model. These are good at allowing for the noise in day to day observations, extracting the trend and projecting it forward.
* Our regression models are designed to exploit this, using the daily statistics released by PHE and NHSE. These strongly suggest that the tide has turned and that taking one day with the next, the national figures for deaths from this virus will now fall back noticeably, easing the pressure on the NHS and its staff.
* There is still a huge range of uncertainty associated with any forecast. The model is currently predicting a total of 113,000 admissions to UK hospitals by the end of April and that 19,000 people will die from the virus in English hospitals by then. There is a 1 in 20 chance that the mortality figures could flatten out more quickly, with around 1,000 more deaths occurring by the end of April. However, there is the same risk that this figure continues to mount, rising to a total of 24,000 by the end of the month. On current trends, the number of deaths in the UK is likely to be 10% higher than the number in England.
* Longer term, the impact of the virus will depend critically upon the likely relaxation of the current government strategy of suppression.

## Introduction

The UK has a great deal of expertise in modelling epidemics. Research teams at Imperial College, Oxford and UCL have developed large models that can be adapted to represent the spread of a new virus like Covid-19.

These models have parameters that represent the degree of contagion, morbidity and ultimately mortality associated with any disease. They are being used to forecast the evolution of the current epidemic and advise the government on the likely effect of interventions like the current lock-down. In these respects, they resemble the large-scale econometric models used by the Office for Budget Responsibility and H.M. Treasury to make economic forecasts and estimate the effect of changes in government economic policy.

However, large models can miss important links, especially when confronted with ‘black swan’ events like a financial crisis or the outbreak of an unknown virus like Covid-19. They are difficult to adapt to such developments and even to update in the light of new data releases from the Office for National Statistics or Public Health England.

Econometricians have developed a variety of simple methods of checking the properties of these models and matching them better to the data. For example, the work of Box and Jenkins (1976) on time series led modelers to regard physical, economic, biological and other large complex systems as a black box, generating data that were best represented in terms of amorphous statistical models. The more recent vector autoregressive model methodology (Sims (1980)) offers a more flexible way of doing this. These approaches are routinely used by City and other analysts to make their economic and financial forecasts. The Bank of England has a suite of economic models used for different tasks, ranging from large scale models for simulating the effects of policy changes and financial shocks to smaller scale models used in economic and financial forecasting.

Similarly, epidemiologists have a range of different ways of modelling epidemics. Many use the logistic model to provide a simple representation of the cumulative number of infections, hospital admissions and deaths caused by an epidemic. To do this they fit a logistic curve to these series. Remarkably, this model accurately predicted the evolution of the Covid-19 outbreak in Wuhan, China (Batista (2020)). However, there are well-known problems with any procedure that fits curves to series that can grow exponentially. In this paper, we show that these problems can be handled using techniques developed for modelling non-stationery economic data, like growth and inflation. We then apply these techniques to model the daily data issued by Public Health England (PHE) and NHS England (NHSE).

These time-series models are relatively simple. They can be updated quickly to take account of the latest data and forecast the spread of the virus through the community. As in any forecasting context, projections need to be updated rapidly in the light of any forecasting errors. These models can distinguish the trend from misleading day-to-day movements and then use this to identify likely turning points and offer an estimate of the final total. They also provide estimates of the uncertainty that inevitably surrounds any forecast at the moment. Although we do not exploit this advantage in the present paper, they also provide useful statistics that summarize the virulence, morbidity and mortality rates in countries with different demographic and other characteristics, allowing the effects of different containment strategies to be estimated.

This paper uses this econometric approach to track the path of the Covid-19 outbreak in the UK and predict its likely evolution. We now have enough data to say, with a reasonable degree of confidence, that the corner has been turned. We focus on the two headline statistics, the daily data released by PHE for identified infections and hospital deaths. Our first model represents the likely daily number of new infections in terms of variables that represent the fractions of the population that on the one hand are likely to have been exposed to the disease and hopefully less susceptible and those that have not and remain susceptible.

The regression equation tracks these data nicely, as shown by Figure 1. The continuous line shows the way this tracks the cumulative number of cases represented by the dots. The continuation shows the path of infections predicted by this equation. We project the latest data forward dynamically given the estimated parameter values assuming that the equation tracks perfectly. This gives an estimate of nearly 113, 000 for the final number of infections (as of 15 April 2020). According to this model, we passed the half way stage of 56, 500 in terms of infections at the beginning of April. This is the ‘point of inflection’ in the jargon, which is the peak of the ‘bell curve’ for new cases, shown by the dotted line.

**Figure 1.**
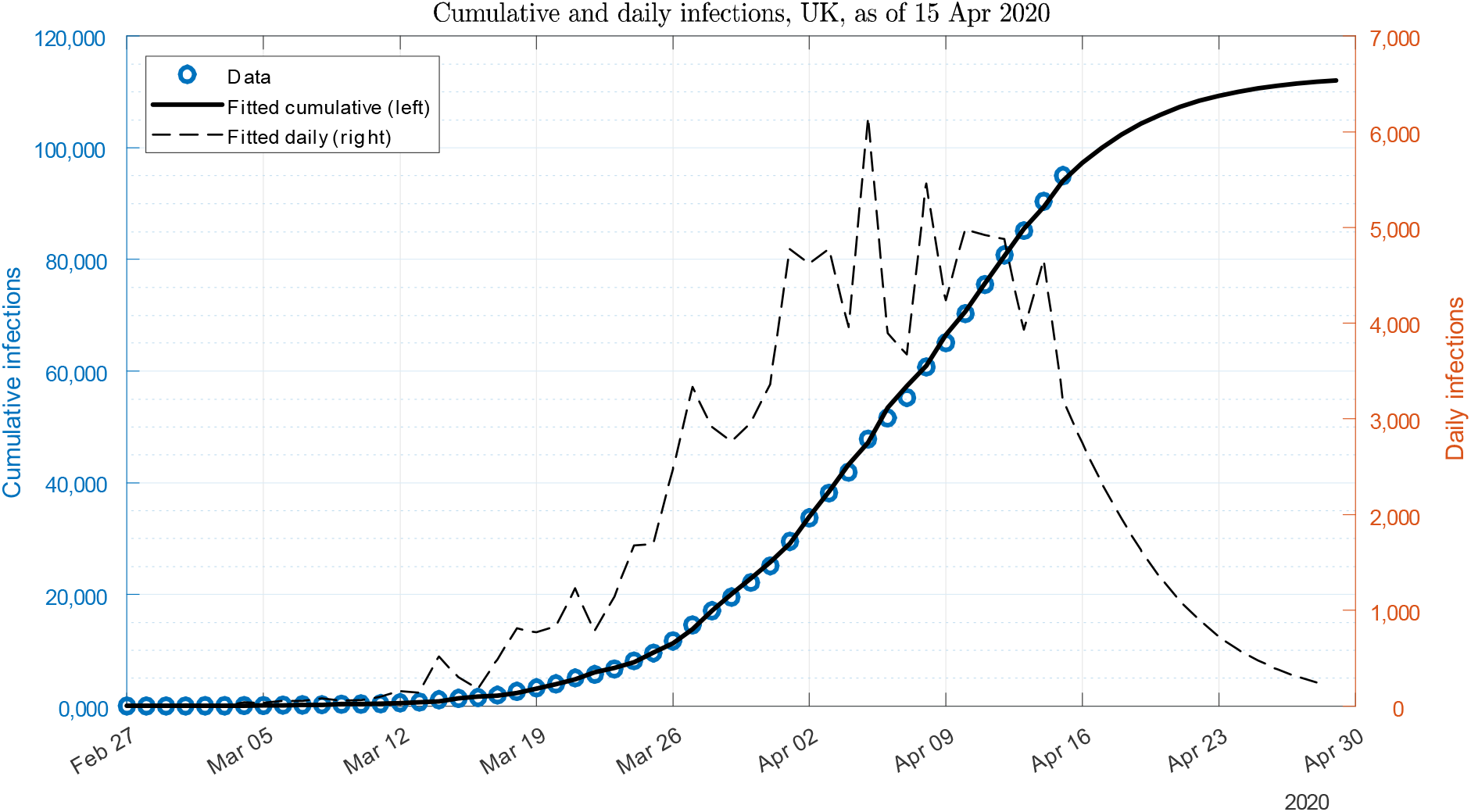

Our second model represents the daily mortality statistics. This sample, though smaller, represents harder data than the infections data and is obviously of crucial importance. The regression equation is depicted in Figure 2, with an estimate of the final number of deaths put at about 15, 500. This estimate is less well determined than in the case of infections, reflecting the shorter data sample. But with almost 13, 000 hospital deaths recorded by 15*th* April, it suggests that we have turned this corner too. Thus, in the same way that the nights gradually start to shorten in January, we should now see the terrible figures for mortality begin to fall, gradually at first but then hopefully gaining pace.

**Figure 2.**
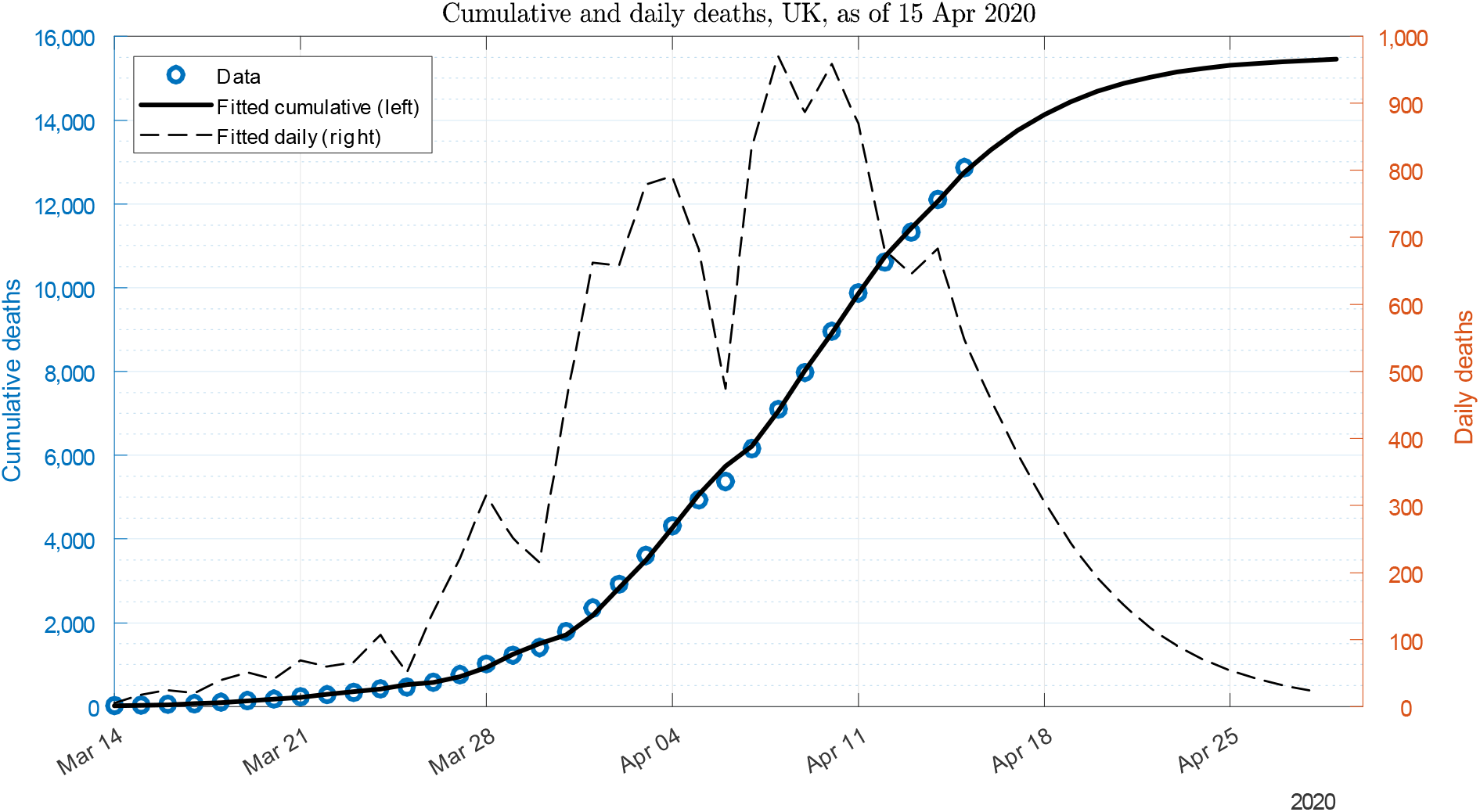

Like economic forecasts, these projections are subject to a considerable degree of uncertainty. This can be assessed relatively easily in these simple models using stochastic simulations, as described in Section 6. Naturally, this uncertainty builds up with the horizon of the forecast, as the effect of simulated errors cumulates over time. Figures 3 and 4 show the results for the cumulative and daily number of deaths, respectively. This clearly illustrates the range of uncertainty still associated with any forecast at this stage of the epidemic. The 90% confidence band in Figure 3 shows that there is a 1 in 20 chance that this number could level out with around 1, 000 more deaths by the end of this month. But there is an equal chance of these figures continuing to mount until the end of the month, rising to a total of 18, 000. After that, the impact of the virus will depend critically upon the likely relaxation of the current government strategy of suppression.

**Figure 3.**
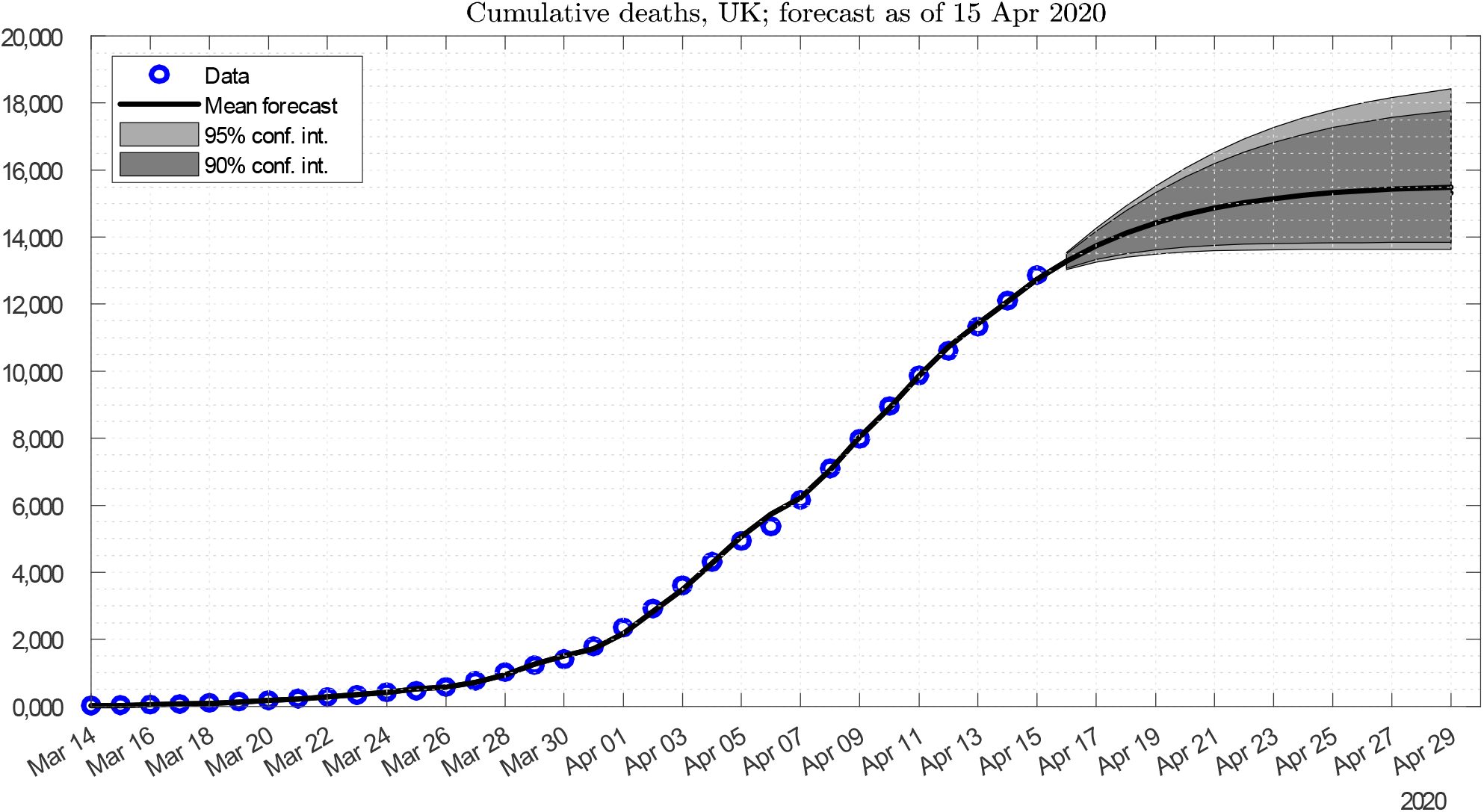

**Figure 4.**
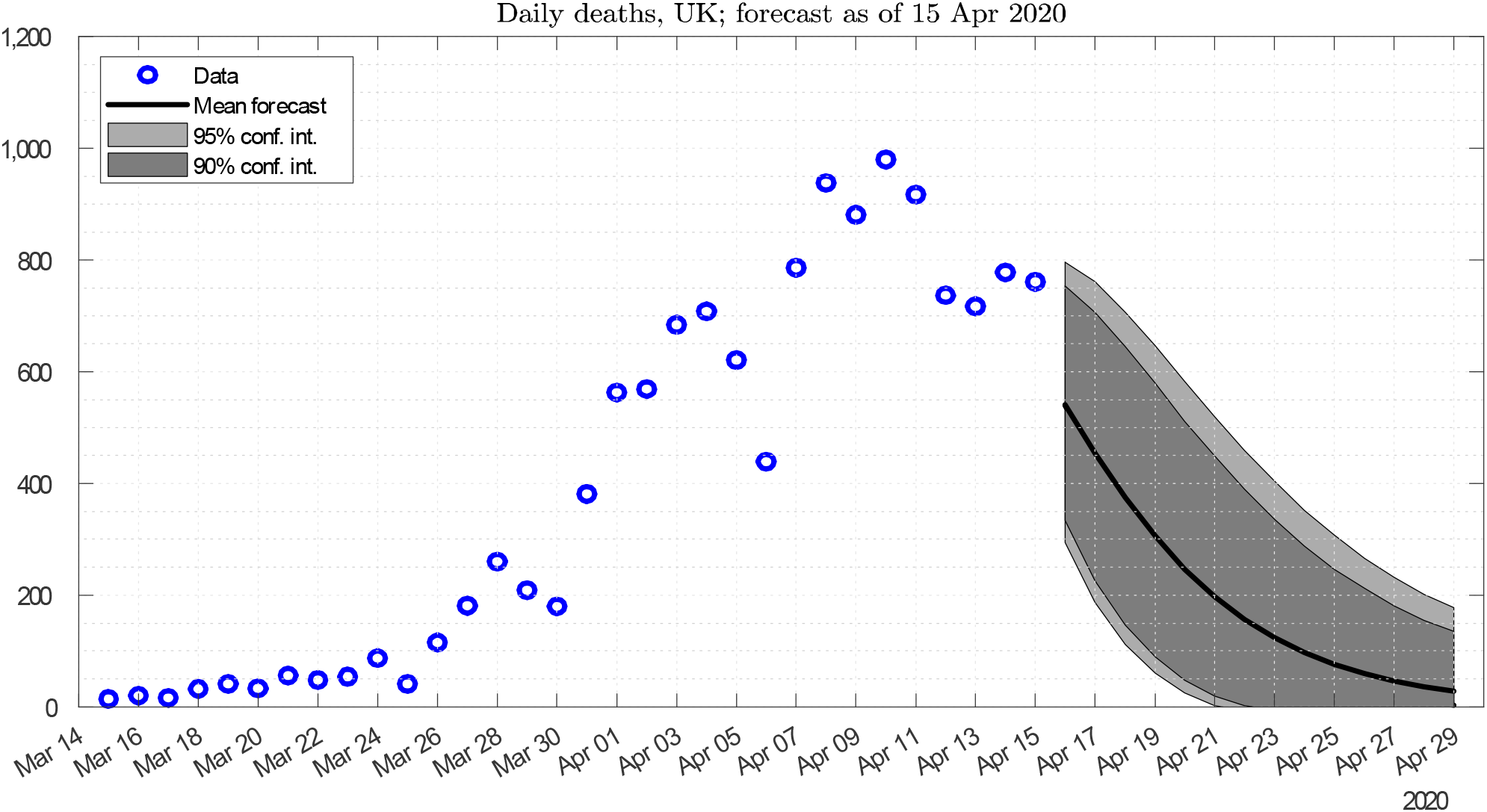

### Caveats

An econometric model is only as good as the data upon which it is based. We need to rely on the PHE statistics despite their drawbacks. The infection figure only counts those who register positive in the swab test, which has until recently focused on hospital admissions. It is likely that this is the tip of a very large iceberg. Indeed, this is currently a subject of controversy between the large scale modelling groups, showing that it is not only economists that are prone to disagreement.

More worrying still, the PHE mortality data only cover hospital deaths and are subject to a long reporting lag. They announce the number of hospital deaths reported to it the previous day. Because relatively few deaths are reported to it on the day they occur, these announcements mainly comprise deaths in previous days. We have explored this issue using another data set released by NHS England.^1^ This takes the deaths reported in England and arranges them by the date of death rather than the date of announcement. This shows that the reporting lag is very long, revealing that at any one time, there has been a large backlog of unreported deaths in England, which understate the running total and artificially inflate the figures announced later. Our estimates, described in Appendix 1, suggest that unless these reporting delays have been significantly reduced, there was a staggering backlog of 4, 800 deaths that had occurred but not yet been reported by 14^*th*^ April. Fortunately it seems that this lag structure is relatively stable, allowing adjustments to the NHSE data that give a good indicator of deaths as they occur. This research looks very promising and is reported in Section 5.

Finally, we would stress that this research is in progress and this discussion paper is not peer reviewed. A new forecast methodology can only be assessed in terms of its track record over time. To help in this assessment, our daily short-term forecasts of the coronavirus mortality rates for the UK are available on our website: https://sites.google.com/york.ac.uk/adam-golinski/coronametrics.

## 1 The logistic process

The basic logistic equation is nicely explained for example in Batista (2020). His equation (1) is:

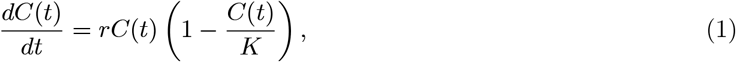

where: *C* is the accumulated number of cases, *K* the final epidemic size and *r* ≥ 0 the propagation or infection rate.

To understand this equation, we can think of infections as being generated by random encounters between Susceptible people (*S*, who have not yet had the disease) and Infectious people (*I*, that are still contagious but not self-isolating). This leaves the rest of the initial population as Removed (*R*), who have already had the infection and have either recovered or died. This is the model of Kermack, McKendrick, and Walker (1927) (originally published in 1927 and republished in 1991). It works like a matching model for new hires in the labour market (which result from employers randomly meeting those looking for work).

Initially, with *C*(0) cases observed when the outbreak is detected, 100% of them will be Infectious and will infect Susceptible people at the rate r per unit of time (*dt*) causing *dC*(0) = *rC*(0)dt new cases. Thus initially, the disease will will spread exponentially, a word which we hear a lot of in common parlance these days. Mathematically this is a very simple differential equation: dC(t) = rC(t)dt. However, as people recover (or die) the probability that an Infected person will meet a Susceptible one declines and amounts to (1 − *C*(*t*)/*K*)), resulting in *rC*(*t*)(1 − *C*(*t*)/*K*)*dt* new cases per unit of time as shown by the equation. This equation has both positive (i.e. Infectious *rC*_*t*-l_) and negative (Removed 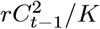) feedback terms. In this deterministic setting *C* will rise inexorably until *C* = *K*.^2^

This is another first-order ordinary differential equation, which we can solve for the path followed by the *accumulated* number of cases *C*. This is the well-known logistic level equation familiar from the charts that we see on television:

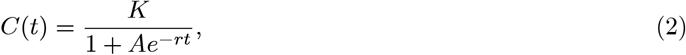

where: *A* = *K*/*C*(0) 1 (see for example Batista (2020), equation (2)). The number of new *daily cases dC*(*t*) is technically the derivative of this function, the figure with the bell shape in Figure 1 that the government is trying to flatten to save the NHS.

The parameters *K* and *r* can be estimated by adding an error term and fitting this to daily or other time series data. However, there are at least two issues with this procedure. First, there are well-known econometric and biometric issues with fitting equations like (2) to level data, which mean we should be working with differential equations like (1). We pick up this point in Section 2 below.

Second, what accounts for the error term: is it external influences, randomness or indeed unrealistic model assumptions? This is critical to the likely outcome. There are several possibilities. The most optimistic is that we eradicate the disease, as we did with smallpox. Next best, we might suppress the disease at a low level as has happened in Wuhan. At the other extreme, it spreads until almost everyone has been infected and is thus immune (the limit *K* is then the initial size of the population).

Either way, *C* can go up but never fall. However, influenza can mutate in a way that having had it once, people can get it a second time. So *rC*/*K*, the fraction of the population that is Infected, can go up and down. That gives a stochastic *equilibrium* or steady state described by a beta distribution, which models the fraction of those Infected in a population as it varies between 0 and *K*. However, that involves longer-term issues that are not our immediate concern. This paper is focussed on modelling the first wave of the coronavirus epidemic in the UK, which we are living through now. The next section is inevitably technical, but the intuition is simple: *C*(*t*) converges monotonically on *K* and then stays there, at least until the next outbreak.

## 2 A stochastic model

If we add a Gaussian random error term *σdz* to (1) this gives the Stochastic Logistic or Stochastic Verhulst diffusion:

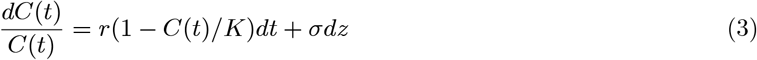

This model is extensively used to model the growth of variables like population and GDP (Merton (1975)). Because the volatility term a-C(t) vanishes as *C*(*t*) tends to zero, it has a lower bound at zero. It has a Gamma distribution in steady state provided that 2*r/σ*^2^ > 1. However, it is unsuitable for modelling the fraction of the population affected by a virus since this is bounded between zero and *K*, while *GDP* and the population itself have an infinite upper bound.

To give the model an upper bound of unity, we also need to ensure that the variance term vanishes as *C*(*t*)/*K* tends to unity. For example we can use the square root volatility specification of Feller (1959) and used to model the lower bound on interest rates by Cox, Ingersoll, and Ross (1985) and Ahn and Gao (1999):

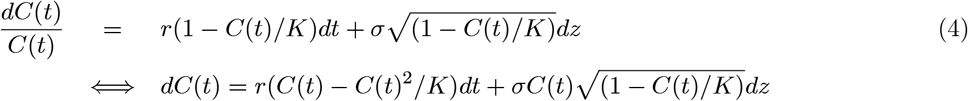

This means that the variance (1 *C*(*t*)/*K*)*C*(*t*) vanishes at both *C*(*t*) = 0 and *C*(*t*)/*K* = 1, ensuring that the process is bounded within the domain [0, *K*]. To test the efficacy of this specification we build an encompassing model that embeds this and other congruent volatility specifications:

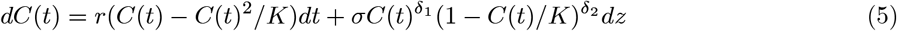

This is a Constant Elasticity of Variance model since the elasticity of the variance 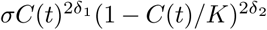 with respect to *C*(*t*) and (1 − *C*(*t*)/*K*) terms is constant (respectively *δ*_l_ and *δ*_2_).

## 3 Modeling the PHE data for infections

To fit this to daily data we then discretize (5) and use it to explain the number of new cases identified in any day:

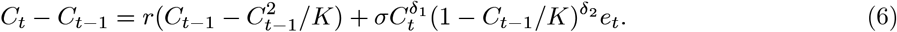

where *C*_*t*_ represents the cumulative number of deaths at time *t* and *e*_*t*_ is an error term, *e*_*t*_ ∼ *N*(0, 1). Given the estimates of the parameters we can then solve for the path of *C* and extrapolate it forward.

This specification assumes that there is no significant infection or recovery lag between the right hand side positive and negative feedback terms and the number of infections being identified in the daily statistics. However, putting in longer lags on these terms reduces the fit of the regression model. This is likely because the recent data represent a larger sample that also reflects infection passed on from people that are missed by the official statistics. It may reflect other measurement errors in the data. This may also reflect behavioral feedbacks. For example, as *C*_*t*_ grows and people hear about the spread of the virus and its consequences, they seem likely to modify their behavior in a way that mimics the effect of the negative ‘Removed’ feedback term (1 − *C*_*t*-l_/*K*), reducing the *K* parameter representing the final total and perhaps speeding up the lags.

We start the estimation sample from 27 February, when the total number of infected cases exceeded 15, since we noted that including the sample by the early period when only a few cases were recorded introduces a bias in the estimation results. Initial experiments showed that the best volatility model had optimal elasticities close to *δ* _l_ = *δ* _2_ = 0.75. Imposing these rounded values gave the result:

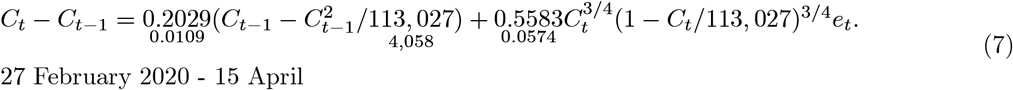

Standard errors are reported in small typeface. The estimate *K* = 113, 027 is reasonably well determined and tells us that according to this model, we are around the half way stage of *K*/2 = 56, 513. This is the point of inflection in Figure 1, which is the peak of the bell curve for new cases. Thus, we should now see the number of new infections start to fall, taking one day with another.

## 4 Modeling the PHE announcements of hospital deaths

Arguably a more important question is: when will we see the mortality statistics peak and at what level? Strictly speaking this requires a different kind of model, since there should be a simple linear relationship between the number of deaths and the number of infections in earlier days. However, the PHE infections statistics record those who register positive in the swab test, which were initially targeted on hospital admissions. It is very likely that this is the tip of a very large iceberg, a subject of controversy as noted in the introduction. In this sense, mortality statistics could represent harder data than infections statistics.

Instead, we can extend the reasoning of the logistic infections model to track deaths instead. Suppose that deaths represent a lagged fraction of the true number of infections. Substituting this into the logistic dynamics (6) gives the model:

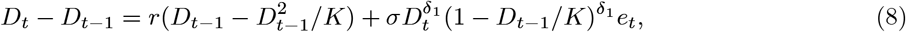

where *D*_*t*_ represents the cumulative number of deaths at time *t* and *K* is now the model parameter representing the final total. Since the mortality record due to Covid-19 is shorter than the total number of infections, we decrease the initial threshold to 10 cases and start the sample from 14 March. In this case the best volatility model had optimal elasticities close to *δ* _l_ = *δ* _2_ = 0.5, giving a square root model. Imposing these rounded values gave the regression result:

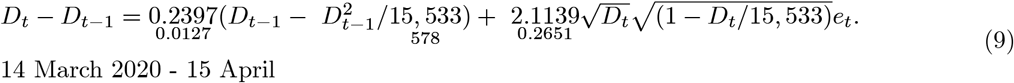

The estimate *K* = 15, 533 is less well determined than in the case of infections, reflecting the shorter data sample.

## 5 Modeling the NHS England date of death data

The PHE data analyzed in the previous section give the number of hospital deaths reported to it the previous day. Because relatively few deaths are reported to it on the day they occur, these mainly comprise deaths in previous days. This is apparent from another data set released by NHS England. This takes the deaths reported in England and arranges them by the date of death rather than the date of announcement. So for example, if we take the total of 778 UK deaths announced on 14^*th*^ April, 744 occurred in England, but only 122 of these occurred on 13th April. Another 319 occurred on 12^*th*^, 132 on the 11^*th*^, 63 on 10^*th*^ and the remaining 108 on previous days. This breakdown of the announcement figures is fairly typical and reveals that at any one time, there has been a large backlog of unreported deaths in England, which inflate the figures announced later. We estimate that on 13^*th*^ April there were a staggering 4, 800 deaths that had occurred but not yet been reported. Our methodology is based on our analysis of the reporting lags seen between March 31 and April 13 and is explained in Appendix 1.

We initially undertook this exercise to assess the scale and structure of the reporting lag. However, we found that this was stable enough from day to day to allow us to estimate the final number of deaths in any day by suitably adjusting the NHS England daily figures. Their figures for the most recent days can be misleading, since they tend to dip because of the reporting lag. Figure 5 shows the daily allocation published on 14 April for example. However, we can use the analysis of the reporting lags in previous weeks to make an allowance for this, as explained in Appendix 1. This adjusted series is also shown the figure. We exclude the figures for 13 April because the number of deaths reported on the day of death is too small to provide a reliable estimate of the true number. Figure 6 shows this alongside the English data arranged by the date of announcement rather than the date of death. On a typical day, these English figures make up 90% of the UK headline total.

**Figure 5:**
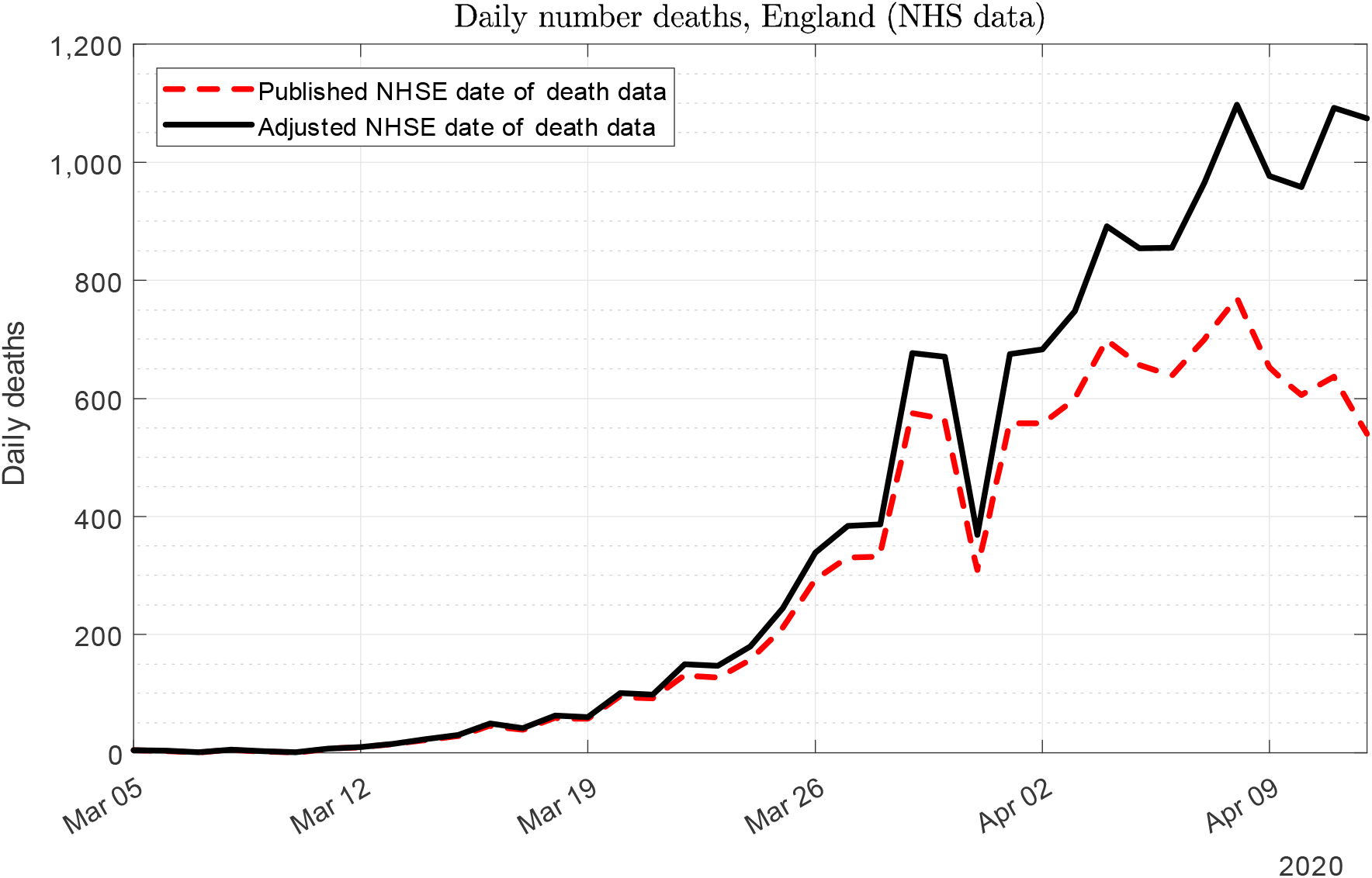
The number of deaths on each day, before and after adjustment.

**Figure 6:**
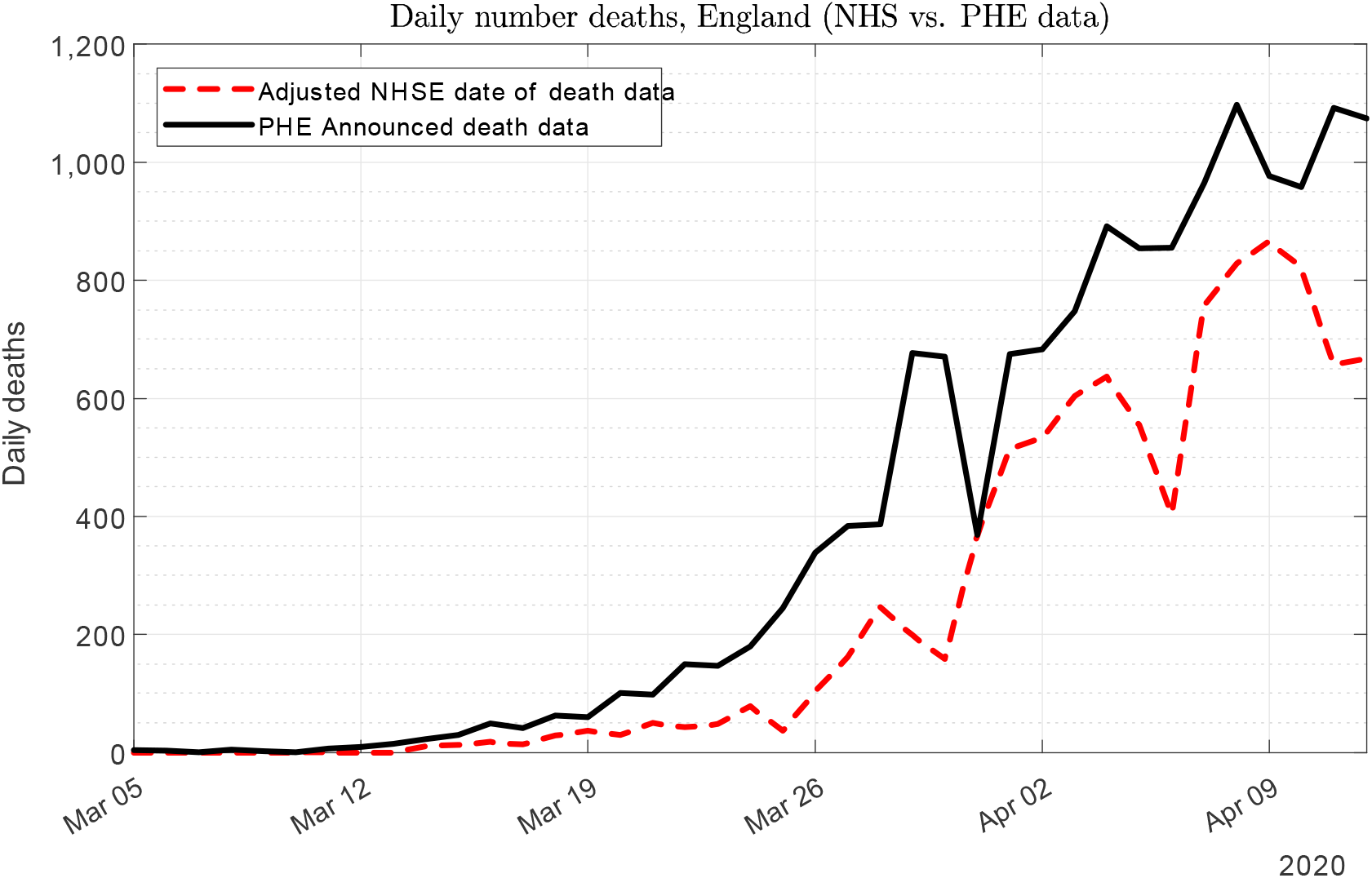
Deaths by day they were announced and the day they occurred.

These adjusted date of death data (*ADD*) offer another way to monitor the effect of the virus on mortality. The regression model give this result:

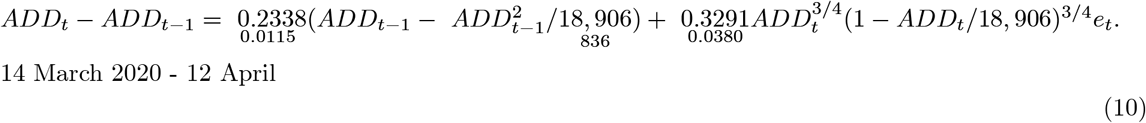

Reflecting the larger number of deaths on this measure, the regression returns a much higher estimate of the final size of the epidemic, *K* = 18, 906, than the announced data set does. This coefficient is better determined statistically, as is the reproduction coefficient: *r* = 0.2338. Figure 7 shows the model fit and extrapolation.

**Figure 7:**
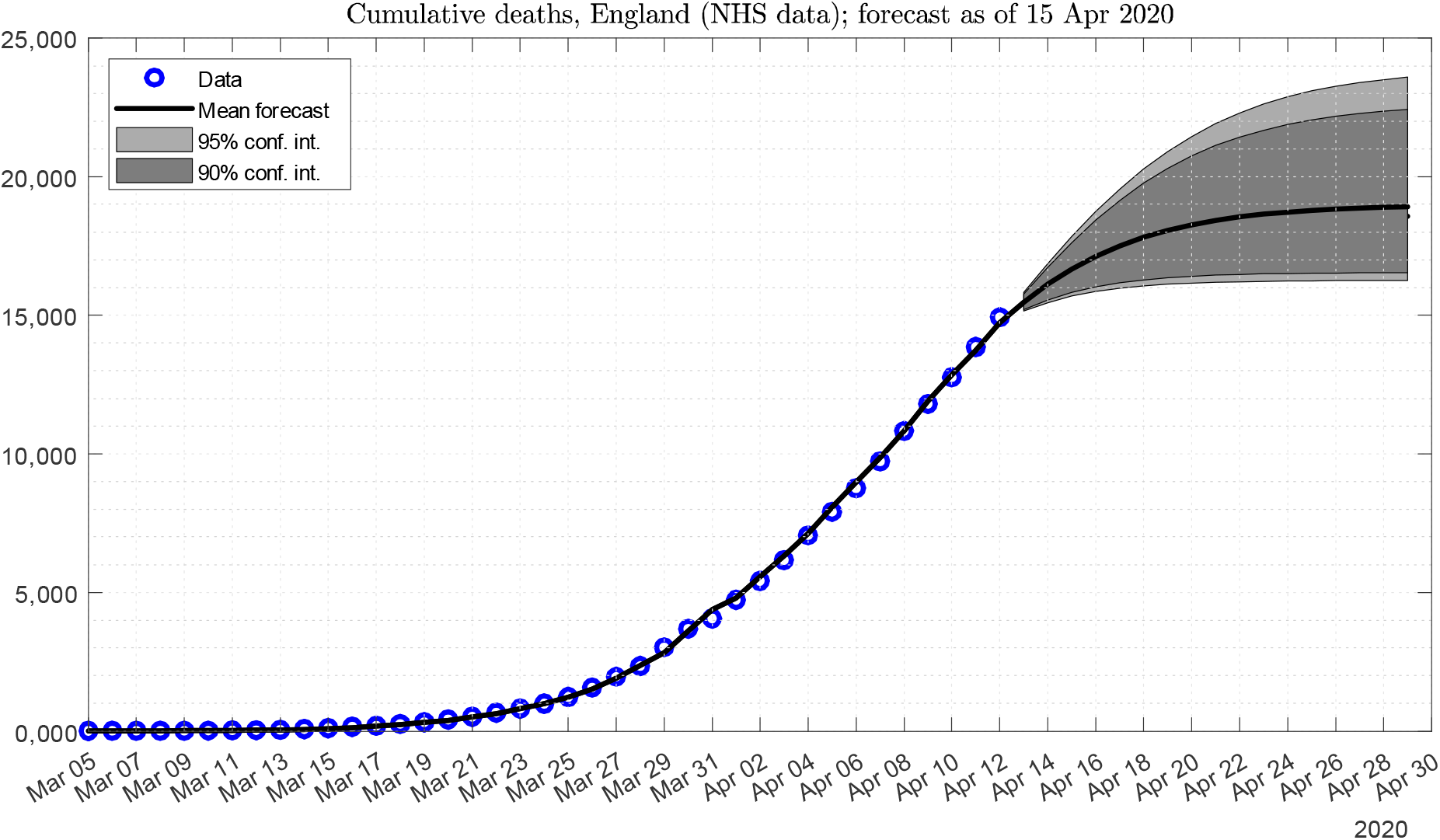
Modeling the adjusted date of death data.

## 6 Allowing for uncertainty

Figures 1 and 2 project these data forward dynamically given the estimated parameter values assuming that the shocks represented by *e*_t_ are zero. However, like economic forecasts, they are subject to uncertainty. This is relatively easy to evaluate in these models using stochastic simulations. Please see Appendix 2 for the technical detail.

There are two sources of uncertainty to evaluate. The first is the additive equation error term _t_, geared up by the volatility term. This source of uncertainty builds up initially with the horizon of the forecast, but eventually declines to zero as the process converges to its limit *K*. The second source of uncertainty concerns the parameters. This is indicated by their estimated standard errors (and covariances). Given these values, a random number generator is used to simulate values of *e*_t_ and errors in the parameter estimates, which are then fed back into the equation dynamics. Repeating this exercise 100, 000 times gives 100, 000 paths for the pattern of infections and deaths due to the virus, which can be used to construct fan charts like Figures 3 and 4, which show the results for the number of deaths. This clearly illustrates the huge range of uncertainty still associated with any forecast at this stage of the epidemic.

## 7 Conclusion

There are many ways of analyzing the economy and the same is true of an epidemic. On the one hand, we need detailed structural models to analyze the likely effects of public health interventions like the current lock-down and on the other we need ways of tracking the progression of the virus and assessing its likely evolution. At the moment for example, some commentators are trying to say where we are on ‘the curve’ and use the experience of other countries that are further along this curve to judge what is likely to happen here. However, as recent experience has shown, it is notoriously difficult to distinguish signal from noise in daily data. It is very easy to make the mistake of thinking that two swallows make a summer. Moreover, the evolution of this virus has differed between countries.

Our approach to this problem, based on our experience of using a range of different approaches to economic forecasting over the years, is that when it comes to short term forecasting, it is very hard to beat a simple time series model. These models are good at allowing for the noise in month to month observation, extracting the trend and projecting it forward. We believe that the same is true of the day to day figures released by PHE and our regression models are designed to exploit this. We are confident that the tide has turned and that taking one day with the next, the national figures for infections, hospital admissions and deaths from this virus will now start to fall back.

## Data Availability

PSE and NHSE data in the public domain

## Appendix 1: Analyzing the NHSE data

The date of death data set released by NHSE is available at: https://www.england.nhs.uk/statistics/statistical-work-areas/covid-19-daily-deaths/

This takes the deaths reported in England and arranges them by the date of death rather than the date of announcement. Two tables are published each day. The first is for the newly announced deaths, which are arranged in terms the date of death. (They are also broken down in terms of age, region and local authority but we focus on the total for England.) The second table cumulates these figures to get the running total for the number of deaths that occurred on each day. These build up over time as deaths are reported and announced.

We treat the first of these tables as revisions, similar to the revisions seen in economic data, which gives different vintages for different release dates. We first express the figure for the revision for each day as a percentage increase on the previous cumulative value for that day. This immediately reveals that the lag is very significant. In particular, the number of deaths reported on the day they occurred is a very unreliable guide to the final total. In the first half of April, it was on average revised up by 237% the next day (vintage 1), 45% the day after (vintage 2) 20% the day after that (vintage 3), and so on and so forth. These average revisions are shown in terms of their vintage in Figure 8.

**Figure 8.**
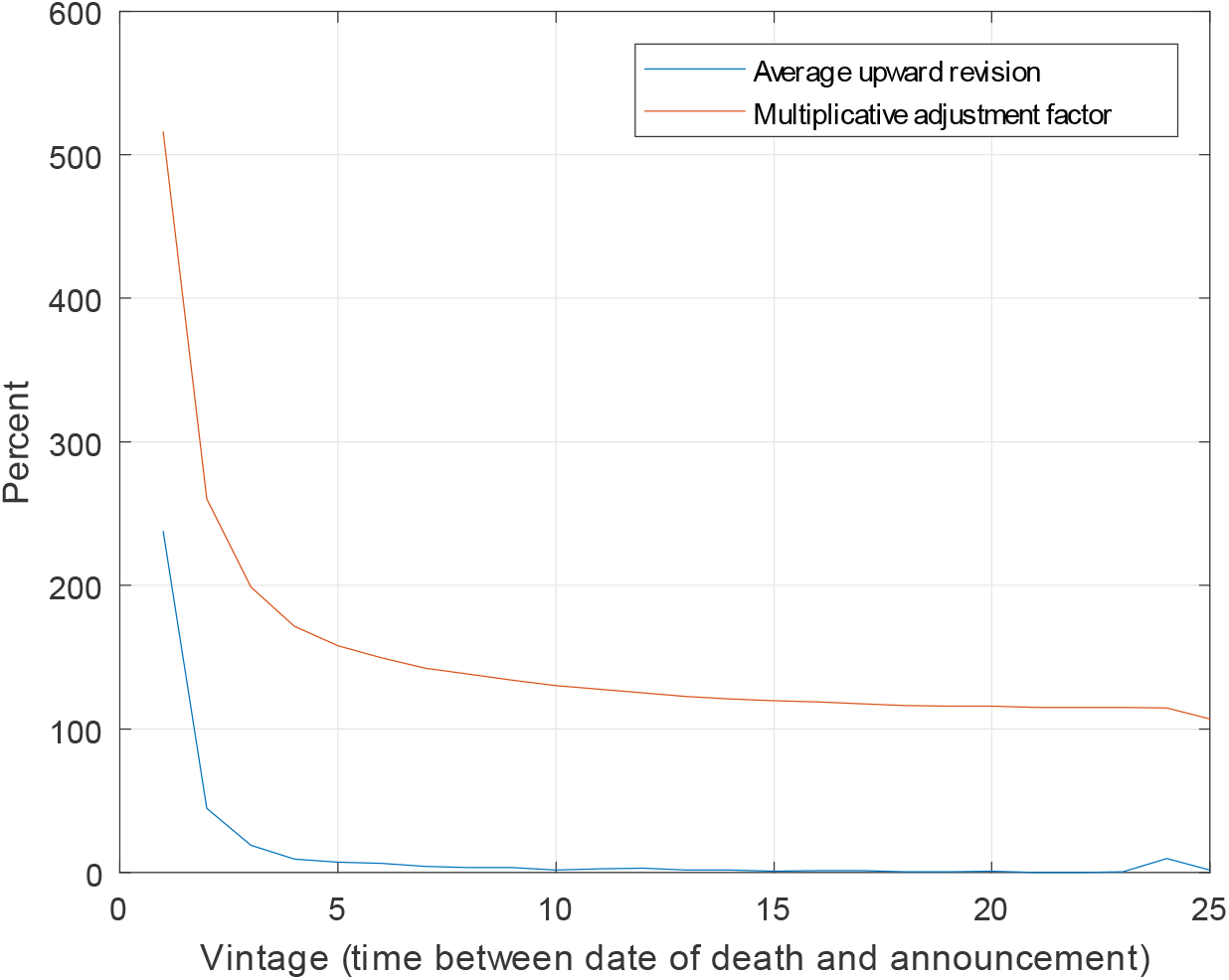
Revisions and adjustment factors by vintage.

We can compound these percentage revisions going backwards in time to get an multiplicative adjustment factor, which, when applied the unadjusted data gives an estimate of the likely final value. These are also depicted in Figure 8. This shows that the number of deaths reported on the day they occurred is on average revised up by a factor of 4.7. After the first revision it is subsequently revised by a factor of 2.3 and so on. These adjustment factors are shown in Figure 8. Applying these factors to the running totals published by NHSE gives our adjusted value. The series for the 13*th* April are depicted in Figure 5. Finally, the cumulative difference between these two series gives an estimate of the number of deaths that have occurred but not been announced. This stood at 4, 800 on 13*th* April. The estimate for 12*th* April is not shown since the adjusted value of the number of deaths reported on the day they occurred is very unreliable. However, we find that this adjustment procedure offers a good indicator after the first and second revision.

Finally, the cumulative difference between the published and adjusted series gives an estimate of the number of deaths that have occurred but not been announced. This stood at 4, 800 on 13th April. A spreadsheet setting out these calculations is available from the authors.

## Appendix 2: Stochastic simulations

We analyse the uncertainty surrounding these forecasts using stochastic simulations. There are two sources of uncertainty to evaluate. The first source of uncertainty concerns the parameters. This is indicated by their estimated standard errors and covariances. To capture the correlation between the parameter estimates, we decompose the variance-covariance matrix of the estimates using the Cholesky decomposition and generate the new set of parameters as

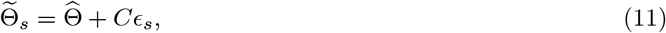

where 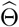 is a 3 x 1 vector of maximum likelihood parameter estimates, *C* is the lower-triangular of the variance matrix of 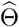 and *ϵ*_s_ is a 3 x 1 vector of standard normal variable generated by a random number generator.

The second source of uncertainty comes from the additive equation error term _t_, geared up by the volatility term. This uncertainty builds up initially with the horizon of the forecast, but eventually declines to zero as the process converges to its limit *K*. Given the (randomized) parameter values, a random number generator of a standard normal variable is used to simulate values of *e*_t_, which are then fed back into the dynamics equation (8).

The inconvenient feature of this simulation approach is that as the process approaches the steady-state *K*, the volatility of the shocks dies out, effectively leaving us only with the parameter uncertainty at the sample. For example, consider two extreme scenarios: one when, by chance, all the unexpected shocks are positive and in the other all unexpected shocks are negative. These shocks have only some limited impact on the forecasts in the middle horizon, but eventually in about three weeks the forecasts are exactly the same. It is rather reasonable to expect that e.g. a large realization of the random shock _t_ (which could result from underestimation of the *K* value) should result in an upward revision of the steady-state. Therefore we propose the following recursive method to update the expectations about the steady-state. Assume that we are at time *T*, i.e. the end of the estimation sample. Denote the estimated *K* by *K*_*T*_. The model predicts that, absent random shocks, the process will change by:

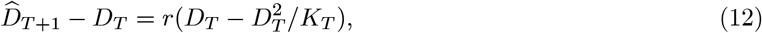

while in fact the (simulated) realization of the process is

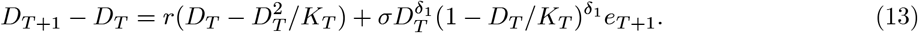

At this point, the unexpected part of the process realization 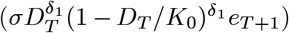 can be considered a contribution to the final value of the steady-state. The value of K that is consistent with this realization is

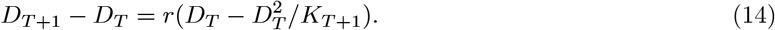

Thus, at time *T* +1 we update of the steady-state as

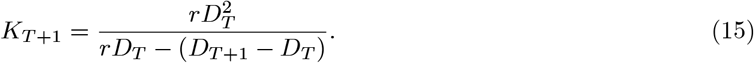

Note that for if the unexpected shock is zero and the value of the realized process is equal to its expectation at time *T* as in (12), then *K*_*T* + l_ = *K*_*T*_. In the next period *T* +2 we repeat this procedure and proceed in this manner until the end of our forecast horizon.

Summarizing, we randomize the parameter values and use them to generate artificial path of the process simulating the random shocks _t_. At each step we update the steady-state *K*. Repeating this procedure 100, 000 times gives 100, 000 paths for the pattern of infections and deaths due to the virus, which can be used to construct fan charts like Figures 3 and 4, which show the results for the number of deaths. This clearly illustrates the huge range of uncertainty still associated with any forecast at this stage of the epidemic.

## Footnotes

1 We are very grateful to Chris Giles, Economics Editor of the *Financial Times* for suggesting we look at this.

2 This is the Verhulst (1838) model. He used it as a deterministic model of population growth, which has similar characteristics, a slow take-off in from a low base, followed by fast growth and finally a slow down as equilibrium is approached.

